# Clinical Evaluation of an AI System for Streamlined Variant Interpretation in Genetic Testing

**DOI:** 10.1101/2025.02.04.25321641

**Authors:** Jiri Ruzicka, Jean-Marie Ravel, Jérôme Audoux, Alexandre Boulat, Julien Thévenon, Kévin Yauy, Marine Dancer, Laure Raymond, Yannis Lombardi, Nicolas Philippe, Michael GB Blum, Nicolas Duforet-Frebourg, Laurent Mesnard

**Affiliations:** SeqOne Genomics, Montpellier, France; Institute of Advanced Biosciences, Univ Grenoble Alpes, CNRS UMR 5309, Grenoble, France; University of Montpellier, LIRMM, CNRS, France; Service de Génétique, Eurofins Biomnis, Lyon, France; Service des Soins intensifs Néphrologiques et Rein Aigu, Hôpital Tenon, Assistance Publique- Hôpitaux de Paris, Paris, France; Centre Maladie rare MAHREA and ERKNET, Hôpital Tenon, Paris, France; INSERM CORAKID, Hopital Tenon, Paris, France

**Author notes:** These authors contributed equally to this work (Jiri Ruzicka, Jean-Marie Ravel).

## Abstract

The growing use of genomic sequencing to diagnose hereditary diseases has increased the interpretive workload for clinical laboratories. Efficient methods are needed to maximize diagnostic yield without overwhelming resources.

We developed DiagAI, an integrative machine-learning system trained to prioritize and sort causal variants in rare diseases. DiagAI integrates Universal Pathogenicity Predictor (UP^2^), a machine-learning model trained to predict ACMG pathogenicity classes, PhenoGenius to match genotype-phenotype interactions and expert features such as inheritance and variant quality.

We retrospectively analyzed 196 diagnosed exomes from a nephrology cohort. To benchmark UP^2^’s performance, we evaluated the ranking of 62 causal missense variants. UP^2^ ranked variants most effectively beyond shortlist sizes of 10 and identified pathogenic variants missed by AlphaMissense. DiagAI identified 94.9% of causal variants in diagnostic exomes with HPO terms, compared to 90.8% without, with median shortlist sizes of 12 and 9 variants, respectively. With HPO terms, 74% of top-ranked variants were diagnostic, versus 42% without, outperforming Exomiser and AI-MARRVEL.

DiagAI produces accurate shortlists that streamline variant interpretation, offering a scalable solution for growing diagnostic volumes.

## Introduction

Hereditary diseases are a significant health concern worldwide, and exome sequencing (ES) and whole genome sequencing (WGS) have become essential for their diagnosis^1,2^. Efficient genomic variant interpretation is a critical step in clinical genomics^3^.

To provide a molecular diagnosis, a large number of variants detected by high throughput sequencing needs to be interpreted. The American College of Medical Genetics and Genomics (ACMG) offers standardized guidelines for variant classification, grouping variants into five categories: pathogenic, likely pathogenic, uncertain significance, likely benign, and benign. Variants of uncertain significance are a major challenge,^4^ that are mostly due to limited available data and insufficient prediction algorithms, although updated recommendations proposed quantitative criteria for pathogenicity classification^5^.

To address these challenges, artificial intelligence (AI) has emerged as a promising solution. Several studies have demonstrated AI’s potential to improve the efficiency of variant interpretation workflows, reduce analysis times, and alleviate the human workload^6–9^ in order to facilitate access to personalized medicine.

We present DiagAI, a machine learning system designed to prioritize the most likely causal variants. DiagAI relies on the prediction of molecular pathogenicity computed by UP^2^ classifier and expert knowledge rules to comprehensively analyze genomics data including family transmission. Furthermore, when phenotypic information coded with Human Phenotype Ontology (HPO)^11^ terms is available, DiagAI upweights genes associated with reported phenotypes to enhance diagnostic precision.

To evaluate the performance of DiagAI, we conducted a retrospective analysis of 966 exomes from patients admitted to an adult nephrology unit. DiagAI successfully shortlisted diagnostic variants in 94.9% of cases when phenotypic information, encoded with HPO terms, was included. In comparison, the success rate was 90.8% when HPO terms were not included.

## Materials and Methods

### Study design

To validate DiagAI, we conducted a retrospective analysis from exome sequencing (ES) data generated from adult participants (n=966) with nephropathy of unknown origin, sequenced from March 2018 to July 2022 (Supplemental Table 1). Of these, 196 (24%) were considered positive cases, defined as containing a causal variant previously identified by a geneticist. The remaining 770 (76%) were considered negative cases, where no diagnosis could be established by a geneticist based on the exome sequencing data.

### Exome sequencing

DNA was extracted from peripheral blood using the QIAsymphony DSP DNA Mini Kit on a QIAsymphony instrument following the manufacturer’s (QIAGEN, The Netherlands) guidelines. Library preparation and capture was performed with Twist reagents (Human Comprehensive Exome or Human Exome 2.0 Plus Comprehensive Exome Spike-in, Twist Bioscience, USA, California). Sequencing was performed on the Illumina NovaSeq6000 (Illumina, USA, California) in paired-end mode (2×150 bp reads). Raw data (bcl format) were converted to FASTQ format using BCL Convert. Reads were aligned to the human reference genome (UCSC Genome Browser build hg37) with Burrows-Wheeler Aligner^12^ for maximal exact matches aligner.

Calling was performed with an internal procedure, the GermlineVar pipeline, of SeqOne Genomics (Montpellier, France). The GermlineVar pipeline implements a comprehensive variant detection strategy utilizing multiple variant calling algorithms. The pipeline integrates Freebayes^13^, GATK^14^, GRIDSS^15^, AluMEI (in-house pipeline^16^), and GATK-Mitochondrial pipeline^17^ (versions ≥2.0). Detection sensitivity parameters are optimized according to the analysis type. In panel-based analyses, the Freebayes algorithm is configured to detect variants with allele frequencies ≥5%, while exome analysis maintains a more stringent threshold of ≥10% for SNVs. Notably, GRIDSS, AluMEI, and GATK operate independently of variant allele frequency thresholds for small variant detection, allowing for maximum sensitivity in structural variant identification.

### Universal Pathogenicity Predictor (UP^2^)

The precise assessment of genomic variant pathogenicity is a fundamental prerequisite for the molecular diagnosis of rare diseases. UP^2^ is a machine learning model using Gradient Boosting Decision Tree algorithm, implemented by the Python package XGBoost^18^, designed to predict molecular pathogenicity of genomic variants. This prediction is based on 74 features derived from various evidence sources (see below) and linked to the ACMG criteria^19^. The classifier assigns a probability to each of the five ACMG classes for a given variant. These five scores are then aggregated into a final UP^2^ score.

The dataset used for training was extracted from ClinVar version 04-2024^10^ and transformed into a standard VCF file using the in-house open-source clinvcf package (https://github.com/SeqOne/clinvcf)^20^. Variants with conflicting interpretations of pathogenicity were re-annotated with removal of outlier submissions falling outside of the inter-quartile range^20^. Variants that stayed conflicting after the reannotation process were excluded. Only single nucleotide variants (SNVs) and short indels were included. Biological annotations of variants were sourced from gnomAD v4.1^21^, VEP predictions version 107^22^ using RefSeq transcripts, dbNSFP^23^ version 4.3, dbscSNV^24^ version 1.1, CI-SpliceAI^25^ v1.1.1, RMSK database^26^ version 210903.

The dataset included 2.7 millions variants from ClinVar^10^, labeled according to the five ACMG pathogenicity categories^19^. It was split into a training set of 2.6 millions of variants, 95% of the total dataset, to train the model using default hyperparameters, and a validation set of 130 000 variants, 5% of the total dataset, to validate the model.

Data augmentation was then used to improve ClinVar data. Indeed, supplementary benign variants, either frequent or non-coding, were added in the dataset to better fit a real-world variant distribution.

### UP^2^ Interpretability features

To assess the relative importance of different ACMG criteria in our classification model, we calculated Shapley values for all 74 features used in the molecular pathogenicity score. These Shapley values were then aggregated in order to be mapped to the known ACMG criteria. This approach allowed us to quantify the contribution of each ACMG tag to the molecular UP^2^ score, providing insight into which criteria were most influential in determining variant pathogenicity according to our model. To calculate a Shapley value for each ACMG criterion, we sum the Shapley values of all features associated with that specific criterion.

### DiagAI priorisation algorithm

The clinical interpretation of genomic data in rare diseases requires determining the pathogenicity of identified variants, establishing a link between the patient phenotypes and genes as well as a concordance between family transmission rules. DiagAI is a linear regression mode, implemented by scikit-learn v1.4.1, trained to retrieve the most likely causal variant for each patient.

The prediction is based on (1) the molecular score UP^2^ detailed above ; (2) a phenotypic score based on Phenogenius algorithm^27^ previously developed and available on GitHub (https://github.com/kyauy/PhenoGenius) and (3) score adjustments based on features linked to inheritance pattern coherence and quality scores. The final score is between 0 and 100.

Phenotypic information was incorporated using HPO^11^ terms, with gene prioritization performed by Phenogenius, a tool that leverages gene-HPO term associations from literature-based matrices^27^. Inheritance mode consistency was sourced from PanelApp^28^ version 240807, OMIM^29^ version 240807, and MedGen^30^ version 230331. Variant calling data (DP>= 5, AO>= 2, VAF>= 0.20), quality (base quality phred score>= 20, no PASS filter used), and parental variant data, in cases where trio sequencing was performed, were based on GermlineVar pipeline detailed above.

The dataset used for training and validation was sourced between 20/10/2021 and 07/11/2023 from proprietary data. The dataset comprised 46 millions variants with 678 causal variants from multiple partner genomic centers, diagnosed by certified molecular pathologists. Patients were affected by rare diseases including intellectual disability, cardiopathy and nephropathy. Causal variant was established by dual-reading by two independent pathologists. Multiple causal variant types are represented in the dataset with a majority of missense. Different configurations of analyses (trio or mono, with phenotypes or without phenotypes) were included to avoid an eventual overfitting of a particular diagnostic scenario. Variants were called by SeqOne in-house pipeline. Risk factor variants (such as *APOL1* variants) were excluded. Only single nucleotide variants (SNVs) and short indels (<300bp) were included.

Missing numerical values were imputed to use a linear regression model. Features were imputed as 0 if the given pattern is not present or not applicable, corresponding to the biological standards of the features. Binary features were already distributed between 0 and 1 included. Concerning continuous features, UP² score was rescaled using min-max normalization and PhenoGenius raw score was rescaled using mean normalization.

The dataset was split into a training set of 26 millions of variants, with 397 causal variants from 307 samples used to train the model parameters and a validation set of 20 millions of variants with 281 diags variants from 252 samples used for validation of DiagAI score prediction

### DiagAI shortlist

The prioritization of variants into a condensed shortlist is a first step in genomic interpretation, enabling the focused functional analysis of a limited set of high-probability candidates. To build a shortlist of variants most likely causal, two thresholds were applied: one for the molecular UP^2^ score and another for the DiagAI score. These thresholds were set to optimize the precision and recall. The DiagAI threshold value depends on the presence of HPO terms. A third threshold was applied to discard variants with a frequency above 1.5% in the phenotypically matched cohort. This threshold was based on the frequency, in our cohort, of the most frequent causal variant.

### Evaluation of performance

We compared the performance of UP^2^ score to rank missense diagnostic variants to the following methods: CADD^31^, REVEL^32^ and AlphaMissense^33^. The thresholds used are taken from the original publications of AlphaMissense^33^ (0.35, 0.55) and REVEL^32,34^ (0.29, 0.644). We focused on missense variants, for which predictive algorithms are key to ACMG classification. To ensure the fairness of our evaluation, particularly given that our UP^2^ model is trained on ClinVar data, we excluded any diagnostic variants previously cataloged in ClinVar.

To benchmark the performance of DiagAI, we compared its ranking performance to AI-MARRVEL and Exomiser (v13), which prioritizes genes or variants by leveraging information on variant frequency, predicted pathogenicity, inheritance modes, and gene-phenotype association^6,35^.

Exomiser13 was run using default pathogenicity sources MVP and REVEL, and failedVariantFilter, inheritanceFilter, frequencyFilter and pathogenicityFilter with the keepNonPathogenic option set to true. After filtering the OmimPrioritizer and hiPhivePrioritizer steps were used. AI-MARRVEL ran with the lite version, and no access to the HGMD resource. Because DiagAI uses in its scoring a filter profile based on quality, variant allele frequency, depth and number of observed alternate alleles, we applied the same filtering prior to the run of Exomiser13 and AI-MARRVEL. We evaluated the rankings at the gene level. For Exomiser13 we used the gene ranks from the json output file. For both DiagAI and AI-MARRVEL genes were ranked based on the highest-priority variant among all their associated variants.

## Results

### Comparison between UP^2^, CADD, REVEL AND AlphaMissense

We compared the ranking provided by UP^2^ to the ones provided by AlphaMissense, CADD and REVEL for causal missense variants from our cohort that were not reported in ClinVar (62 variants, Figure 1). For the shorter shortlist (<=10), REVEL outperformed other tools in prioritizing causal missense variants: it ranked the causal variant first in 12.90% of cases and within the top 10 in 32.26%. In comparison, the top 1 / top 10 inclusion rates were 0% / 27.42% for UP^2^, 0% / 1.61% for AlphaMissense, and 0% / 1.61% for CADD. For larger variant lists (>10), UP^2^ outperformed other methods and ranked the causal variant within the top-100 shortlist in 87.10% of cases. In comparison, the percentage of top-100 shortlist containing the causal missense variants were 61.29% for REVEL, 19.35% for AlphaMissense, and 11.29% for CADD.

**Figure 1:**
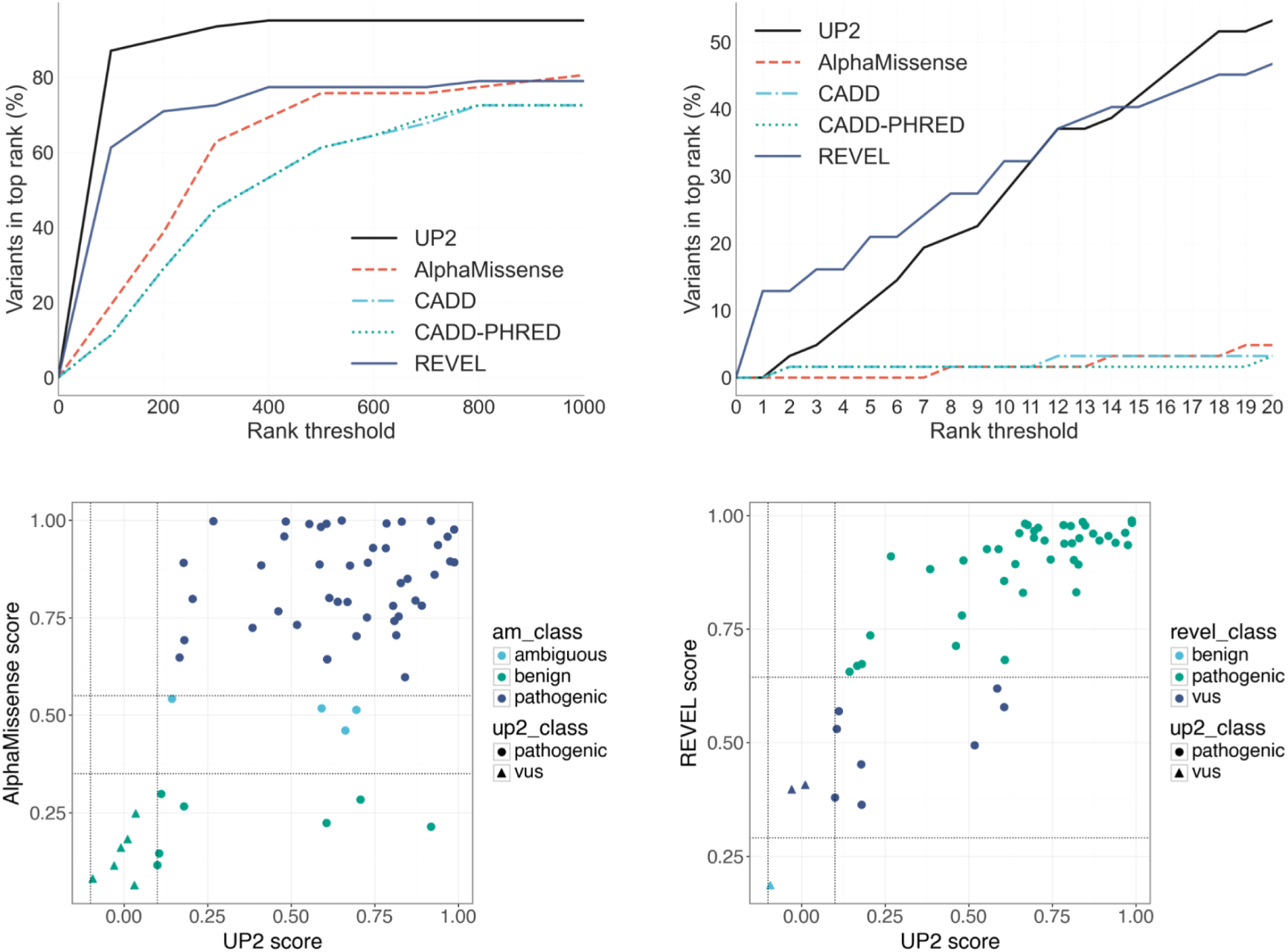
Performance of UP^2^ variant ranking evaluated on 62 missense variants not reported in ClinVar. Top-level panel : percentage of analysis with the diagnostic variant being in the top-rank list of variants (left: top 1000, right: top 20) according to different scoring systems: AlphaMissense, CADD, REVEL and UP^2^. Bottom-level panel : Variants’ scores comparison between AlphaMissense (left) or REVEL (right) and UP^2^ (62 causal missense variants not in ClinVar). am_class : classification predicted by AlphaMissense ; up2_class : classification predicted by UP^2^ ; am_revel: classification predicted by REVEL. Dashlines indicate the threshold of corresponding algorithms. The down right corner of Alphamissense vs UP^2^ comparison corresponds to the variants not detected by AlphaMissense but reported as pathogenic by the UP^2^ scoring system.

Comparing AlphaMissense and UP^2^ scores only, we find strong concordance with 72.6% (45/62) of the variants classified as pathogenic by both methods. However, 7 variants (11.3%) were incorrectly classified as benign by AlphaMissense whereas they were correctly considered as pathogenic by UP^2^ (Figure 1, lower left panel). No variants were considered as pathogenic by AlphaMissense and benign with UP^2^. REVEL and UP^2^ also showed strong concordance, with only one causal variant classified as benign by REVEL and as VUS by UP^2^, and no cases where one method classified a variant as pathogenic and the other as benign. (Figure 1, lower right panel).

### Interpretability of the classifier

We applied our interpretability framework to identify the ACMG criteria most influential in determining the ACMG classifications of our cohort of 196 exomes consisting of 176 unique causal variants. Some variants were indeed shared between exomes. Among the 85 diagnostic variants with ClinVar submissions, features related to the number of pathogenic and benign submissions were the most influential (Figure 2). However, for 13 variants, other ACMG criteria were more impactful, including 6 with PP3/BP4 (in-silico predictors of pathogenicity and benignity), 5 with PVS1 (predicted impact by VEP), and 2 with PM2/BA1 (absent or at extremely low frequency in general population).

**Figure 2.**
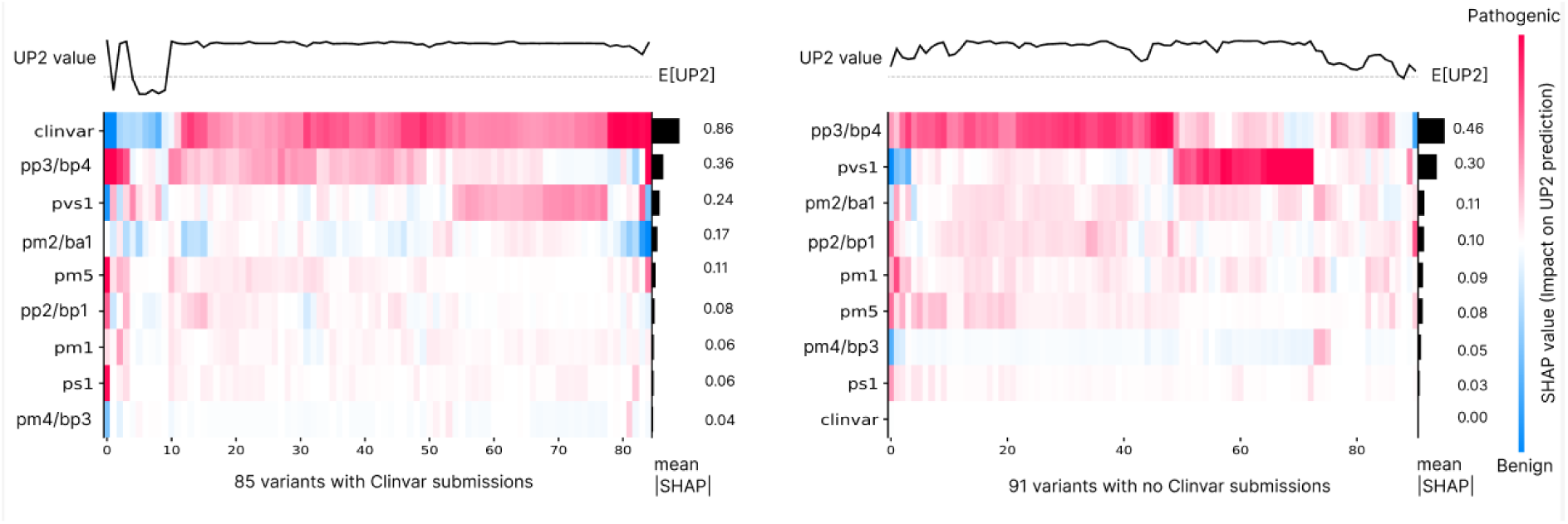
Explicability profile of the 176 diagnostic variants for the Universal Pathogenicity Predictor. The heatmap shows for each variant if the impact of the features related to an ACMG criteria is rather pathogenic (red) or benign (blue). Variants that have ClinVar Submissions (Panel A, left) have UP^2^ predictions that are mostly influenced by the ClinVar submission predictors (number of submissions per ACMG class) with an average absolute SHaP value of .86 for ClinVar predictors. Variants that have no ClinVar Submission (Panel B, right) have UP^2^ predictions that are mostly influenced by features from PP3/BP4 or PVS1 criteria. The right side histogram shows that the average absolute SHaP value is .46 for PP3/BP4 and .30 for PVS1 on the UP^2^ value for variants without ClinVar submission. The top plots show UP^2^ values that are between -1 and 1.

For the 91 diagnostic variants without ClinVar submissions, PP3/BP4 features were instrumental for 66% (60/91) of the variants, while PVS1 features were key for 29% (26/91). The scores for the remaining five variants were primarily determined by a combination of other ACMG features.

### Proportion of Causal Variants Identified in Shortlists

For exomes with a confirmed molecular diagnosis, 94.9% (186/196) of causal variants were contained in the shortlist when HPO terms were used, compared to 90.8% (176/196) when HPO terms were not used. The median shortlist size was 9 variants when HPO terms were not used (min=2, max=44) and 12 variants when HPO terms were used (min=4, max=29).

### Variant ranking

We compared DiagAI’s variant ranking accuracy with and without utilizing HPO-based clinical information (Figure 3) on the 196 exomes with a confirmed diagnostic variant.

**Figure 3.**
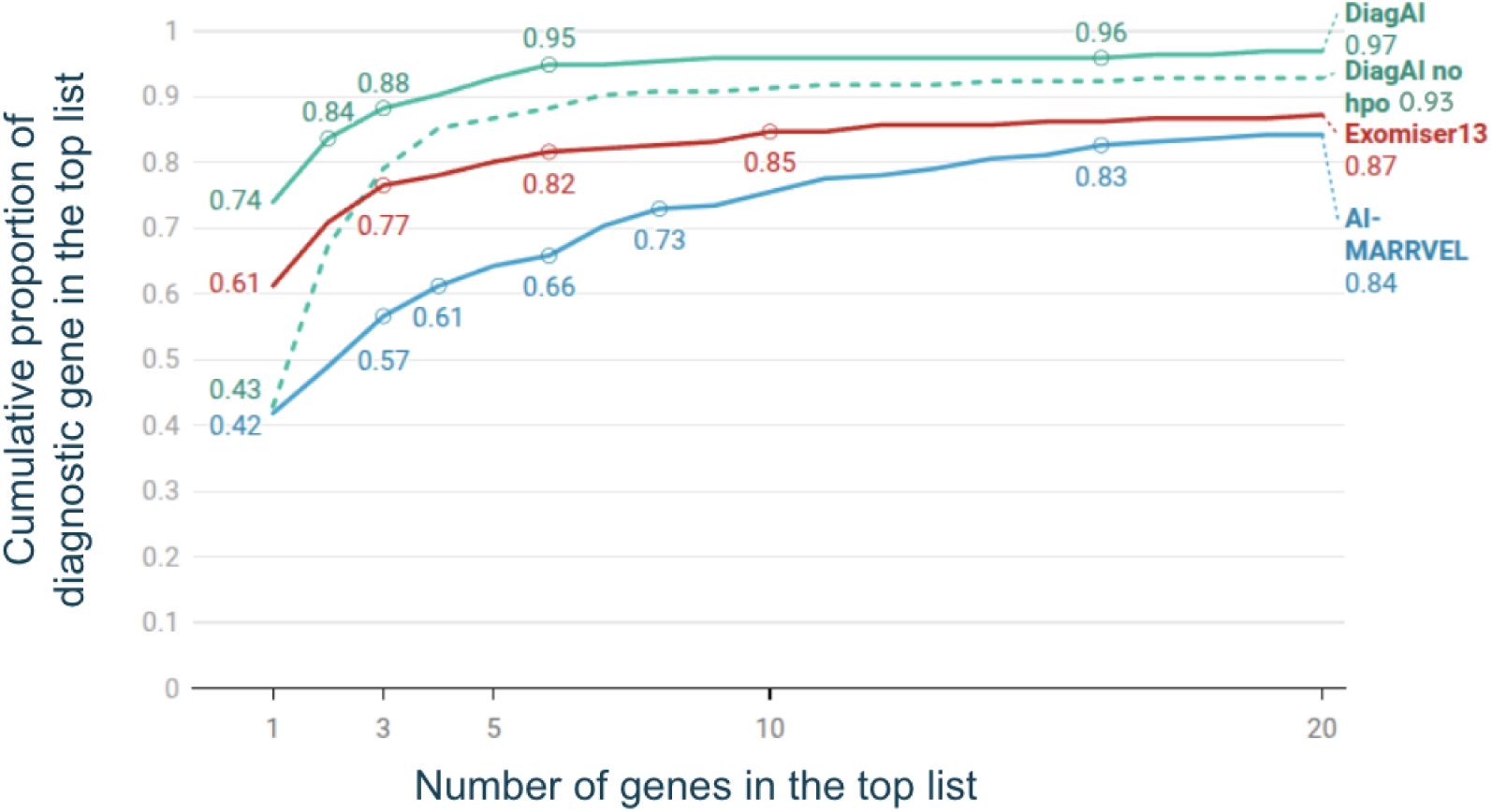
Performance of variant ranking evaluated on the 196 exomes with a confirmed molecular diagnosis. The ranking accuracy of top-ranked genes was assessed using two approaches when evaluating DiagAI: the molecular pathogenicity score alone (no HPO) and a comprehensive score integrating molecular pathogenicity and HPO-based clinical information (with HPO). Variants with a cohort frequency above 1.5% were excluded from the DiagAI ranking. Exomiser v13 and AI-MARRVEL, which also use HPO for gene ranking, were used for comparison.

Incorporating clinical data significantly improved the ranking accuracy for the top-ranked variant. Specifically, 42% of top-ranked variants were diagnostic when HPO terms were not included, whereas this percentage increased to 74% when HPO terms were accounted for. The improvement was less pronounced when considering a larger list of 20 genes, with 93% of diagnostic variants included without HPO terms versus 97% with HPO terms. DiagAI achieved improved ranking performance compared to Exomiser v13 and AI-MARRVEL when HPO terms were provided, and for top-ranked lists of three or more variants when HPO terms were not included (Figure 3).

### Causal variants absent from the shortlists

A total of 10 diagnostic variants were absent from the DiagAI shortlists, despite being considered diagnostic by the geneticists. Of these, five were classified as benign in ClinVar, and three were labeled as variants of uncertain significance (VUS) or had conflicting classifications in ClinVar, with no additional evidence supporting pathogenicity. The remaining two variants were missed by DiagAI due to specific limitations: one was a low-quality variant, and the other was located in a recessive gene without a second heterozygous variant with a sufficiently high DiagAI score to support a composite diagnosis.

## Discussion

Our study demonstrates DiagAI’s effectiveness in prioritizing causal variants in exomes from nephrology patients, analyzed as single cases rather than trios. Depending on the availability of phenotypic data, 90.8 to 94.9% of shortlists contained the causal variant. DiagAI also outperformed Exomiser v13 and AI-MARRVEL in gene-level ranking comparisons.

We also compared UP^2^ to widely used variant scoring tools to assess its utility in clinical variant prioritization, focusing specifically on causal missense variants not reported in ClinVar. REVEL performed well in ranking top variants but failed to capture several true positives as shortlist size increased. In contrast, UP^2^ maintained strong performance across broader ranking ranges, making it more suitable for diagnostic applications where sensitivity is essential. AlphaMissense and CADD showed consistently lower performance, underscoring the added value of UP^2^, and to a lesser extent REVEL, for effective variant prioritization.

DiagAI outperforming open source alternatives is explained by several key features. First, the UP^2^ algorithm was meticulously designed through the selection and analysis of a comprehensive set of features to enhance the model’s ability to identify pathogenic variants. A key contribution lies also in the rigorous data curation process during the ClinVar data treatment, addressing conflicting ClinVar classifications ^20^. Furthermore, we implement data augmentation techniques to mitigate biases, particularly those stemming from ClinVar’s inherent skew toward pathogenic variants, which can distort model performance for example in intergenic regions. Secondly, DiagAI implements a modular two-layer architecture that integrates machine learning models trained on patient cohorts with expert-driven methodologies to capture complex patterns. This approach addresses critical challenges, including the accurate assessment of variant inheritance, notably the interpretation of compound heterozygous variants, as well as the evaluation of data quality and the management of sequencing technology differences. The integrated design not only strengthens diagnostic reliability but also enhances the framework’s adaptability to a wide range of data types, including long-read sequencing (100% precision on a cohort of ten long read whole genome sequences, data not shown).

We examined some of the causal variants missed by the shortlist of DiagAI. We identified one variant in the *PODXL* gene, whose pathogenic effect has been linked to kidney diseases^36^. We also identified a variant in *CFI*, which dysregulates the complement alternative pathway. However, *CFI* variants are challenging to interpret; they are difficult to classify using ACMG criteria^37^, and due to their incomplete penetrance, they are often considered risk factors rather than causative in a Mendelian disease^38^. Additionally, a synonymous variant in *NPHP3* with a splicing effect, not captured by computational approaches, was also overlooked^39^. Overall, these variants were missed by DiagAI but were identified as diagnostic by geneticists thanks to recent research documenting their pathogenicity, combined with expert knowledge of genes involved in kidney diseases.

These cases illustrate the inherent challenges in variant interpretation, particularly for variants with emerging or complex pathogenic mechanisms that are not yet well captured by computational models. While DiagAI improves variant prioritization by leveraging multivariate ACMG evidence tags and machine learning trained on ClinVar ACMG classifications, it remains limited by the available knowledge and data used for training. By integrating diverse evidence sources and phenotypic data encoded with HPO terms, DiagAI enhances variant ranking, but expert review remains essential for capturing novel or particularly challenging cases.

DiagAI contributes to the growing landscape of computational solutions for variant prioritization, joining both open-source and commercial offerings such as AI-MARRVEL, InVitae MOON, Fabric GEM, and the Emedgne software from Illumina^6,8,9,40^. Direct comparisons between these tools are challenging due to differences in their evaluation cohorts. Nonetheless, we found that DiagAI outperformed AI-MARRVEL in gene ranking and demonstrated comparable performance in identifying diagnosable cases, with both tools automatically detecting 50–60% of such cases^6^. For a fair comparison, the Critical Assessment of Genome Interpretation (CAGI) challenge offers a standardized benchmarking framework; however, DiagAI has yet to be evaluated within this context.

DiagAI’s accuracy in variant ranking, particularly when integrating clinical data, highlights its potential to streamline genomic diagnostics by reducing the number of variants requiring manual review. However, the path to full automation remains long, with less than 60% of diagnosed cases detected automatically. These findings suggest that AI-powered tools like DiagAI can significantly reduce the interpretive workload in clinical genomics while maintaining high diagnostic accuracy. This assessment should be evaluated beyond the specific case of nephrology and further tested in the context of whole genome sequencing analysis.

## Data Availability Statement

Data should be requested to Prof. L Mesnard.

## Code Availability

ClinVCF and Phenogenius are open source and available on GitHub https://github.com/SeqOne/clinvcf and https://github.com/kyauy/PhenoGenius.

## Acknowledgments

We would like to thank the technicians, scientists, bioinformaticians, operations staff, and biologists at Eurofins Biomnis for their valuable contributions to the production and interpretation of these data.

## Author Contributions Statement

Conceptualization: M.G.B., N.P., N.D.-F., L.M.; Methodology: J.R., K.Y., N.D.-F., J.T.; Data Curation: J.R., A.B.,; Data Analysis: J.R., N.D.-F.,J.-M.R.; Data Production: L.M., M.D., Y.L., L.R., J.-M.R.; Software: J.R., N.D.-F., J.A., N.P.; Supervision: M.G.B., N. D.-F.; Writing – Original Draft: M.G.B.; Writing – Review and Editing: J.-M.R., M.G.B., L.M., J.R

## Funding

This study was conducted without additional funding.

## Ethics Statement

The cohort has been previously described^2^. The study was approved by an Institutional Review Board (Direction de la Recherche Clinique et de l’Innovation (APHP220461) and the Ethic board of Sorbonne Université (CER-2022-009).

## Competing Interest

J. Ruzicka, J.-M. Ravel, J. Audoux, A. Boulat, K. Yauy, N. Philippe, M. Blum, and N. Duforet-Frebourg are current or former employees of SeqOne Genomics and hold, or have received, stock or stock options from the company. M. Dancer and L. Raymond are employees of Eurofins Biomnis.

## Notes

### Author Declarations

The cohort has been previously described. The study was approved by an Institutional Review Board (Direction de la Recherche Clinique et de l Innovation (APHP220461) and the Ethic board of Sorbonne Université (CER-2022-009).

### Summary of Updates

Improve the manuscript overall with great emphasis on methodology.

## References

1. 1. 100,000 Genomes Pilot on Rare-Disease Diagnosis in Health Care — Preliminary Report. N Engl J Med. 2021;385(20):1868–1880. doi:10.1056/NEJMoa2035790

2. Doreille A, Lombardi Y, Dancer M, et al. Exome-First Strategy in Adult Patients With CKD: A Cohort Study. Kidney Int Rep. 2023;8(3):596–605. doi:10.1016/j.ekir.2022.12.007

3. Austin-Tse CA, Jobanputra V, Perry DL, et al. Best practices for the interpretation and reporting of clinical whole genome sequencing. NPJ Genomic Med. 2022;7(1):27.

4. Amendola LM, Muenzen K, Biesecker LG, et al. Variant Classification Concordance using the ACMG-AMP Variant Interpretation Guidelines across Nine Genomic Implementation Research Studies. Am J Hum Genet. 2020;107(5):932–941. doi:10.1016/j.ajhg.2020.09.011

5. Pejaver V, Byrne AB, Feng BJ, et al. Calibration of computational tools for missense variant pathogenicity classification and ClinGen recommendations for PP3/BP4 criteria. Am J Hum Genet. 2022;109(12):2163–2177.

6. Mao D, Liu C, Wang L, et al. AI-MARRVEL — A Knowledge-Driven AI System for Diagnosing Mendelian Disorders. NEJM AI. 2024;1(5). doi:10.1056/AIoa2300009

7. Owen MJ, Lefebvre S, Hansen C, et al. An automated 13.5 hour system for scalable diagnosis and acute management guidance for genetic diseases. Nat Commun. 2022;13(1):4057.

8. De La Vega FM, Chowdhury S, Moore B, et al. Artificial intelligence enables comprehensive genome interpretation and nomination of candidate diagnoses for rare genetic diseases. Genome Med. 2021;13(1):153. doi:10.1186/s13073-021-00965-0

9. Meng L, Attali R, Talmy T, et al. Evaluation of an automated genome interpretation model for rare disease routinely used in a clinical genetic laboratory. Genet Med. 2023;25(6):100830.

10. Landrum MJ, Chitipiralla S, Brown GR, et al. ClinVar: improvements to accessing data. Nucleic Acids Res. 2020;48(D1):D835–D844. doi:10.1093/nar/gkz972

11. Köhler S, Doelken SC, Mungall CJ, et al. The Human Phenotype Ontology project: linking molecular biology and disease through phenotype data. Nucleic Acids Res. 2014;42(D1):D966–D974. doi:10.1093/nar/gkt1026

12. 12. Li H. Aligning sequence reads, clone sequences and assembly contigs with BWA-MEM. Published online May 26, 2013. doi:10.48550/arXiv.1303.3997

13. 13. Garrison E, Marth G. Haplotype-based variant detection from short-read sequencing. Published online July 20, 2012. doi:10.48550/arXiv.1207.3907

14. 14. Auwera G van der, O’Connor BD. Genomics in the Cloud: Using Docker, GATK, and WDL in Terra. First edition. O’Reilly Media; 2020.

15. Cameron DL, Schröder J, Penington JS, et al. GRIDSS: sensitive and specific genomic rearrangement detection using positional de Bruijn graph assembly. Genome Res. 2017;27(12):2050–2060. doi:10.1101/gr.222109.117

16. Uguen K, Redon S, Rouault K, et al. An unusual diagnosis of alpha-mannosidosis with ocular anomalies: Behind the scenes of a hidden copy number variation. Am J Med Genet A. 2024;194(5):e63532. doi:10.1002/ajmg.a.63532

17. McKenna A, Hanna M, Banks E, et al. The Genome Analysis Toolkit: A MapReduce framework for analyzing next-generation DNA sequencing data. Genome Res. 2010;20(9):1297–1303. doi:10.1101/gr.107524.110

18. Chen T, Guestrin C. XGBoost: A Scalable Tree Boosting System. In: Proceedings of the 22nd ACM SIGKDD International Conference on Knowledge Discovery and Data Mining. ACM; 2016:785–794. doi:10.1145/2939672.2939785

19. Richards S, Aziz N, Bale S, et al. Standards and guidelines for the interpretation of sequence variants: a joint consensus recommendation of the American College of Medical Genetics and Genomics and the Association for Molecular Pathology. Genet Med. 2015;17(5):405–423. doi:10.1038/gim.2015.30

20. Yauy K, Lecoquierre F, Baert-Desurmont S, et al. Genome Alert!: A standardized procedure for genomic variant reinterpretation and automated gene-phenotype reassessment in clinical routine. Genet Med Off J Am Coll Med Genet. 2022;24(6):1316–1327. doi:10.1016/j.gim.2022.02.008

21. Karczewski KJ, Francioli LC, Tiao G, et al. The mutational constraint spectrum quantified from variation in 141,456 humans. Nature. 2020;581(7809):434-443. doi:10.1038/s41586-020-2308-7

22. McLaren W, Gil L, Hunt SE, et al. The Ensembl Variant Effect Predictor. Genome Biol. 2016;17(1):122. doi:10.1186/s13059-016-0974-4

23. Liu X, Li C, Mou C, Dong Y, Tu Y. dbNSFP v4: a comprehensive database of transcript-specific functional predictions and annotations for human nonsynonymous and splice-site SNVs. Genome Med. 2020;12(1):103. doi:10.1186/s13073-020-00803-9

24. Jian X, Boerwinkle E, Liu X. In silico prediction of splice-altering single nucleotide variants in the human genome. Nucleic Acids Res. 2014;42(22):13534–13544. doi:10.1093/nar/gku1206

25. Strauch Y, Lord J, Niranjan M, Baralle D. CI-SpliceAI—Improving machine learning predictions of disease causing splicing variants using curated alternative splice sites. PLOS ONE. 2022;17(6):e0269159. doi:10.1371/journal.pone.0269159

26. Jurka J. Repbase update: a database and an electronic journal of repetitive elements. Trends Genet TIG. 2000;16(9):418–420. doi:10.1016/s0168-9525(00)02093-x

27. Yauy K, Duforet-Frebourg N, Testard Q, et al. Learning phenotypic patterns in genetic disease by symptom interaction modeling. Published online July 31, 2022:2022.07.29.22278181. doi:10.1101/2022.07.29.22278181

28. Martin AR, Williams E, Foulger RE, et al. PanelApp crowdsources expert knowledge to establish consensus diagnostic gene panels. Nat Genet. 2019;51(11):1560–1565. doi:10.1038/s41588-019-0528-2

29. Amberger J, Bocchini CA, Scott AF, Hamosh A. McKusick’s Online Mendelian Inheritance in Man (OMIM). Nucleic Acids Res. 2009;37(Database issue):D793–796. doi:10.1093/nar/gkn665

30. Sayers EW, Beck J, Bolton EE, et al. Database resources of the National Center for Biotechnology Information in 2025. Nucleic Acids Res. 2025;53(D1):D20–D29. doi:10.1093/nar/gkae979

31. Rentzsch P, Witten D, Cooper GM, Shendure J, Kircher M. CADD: predicting the deleteriousness of variants throughout the human genome. Nucleic Acids Res. 2019;47(D1):D886–D894. doi:10.1093/nar/gky1016

32. Ioannidis NM, Rothstein JH, Pejaver V, et al. REVEL: An Ensemble Method for Predicting the Pathogenicity of Rare Missense Variants. Am J Hum Genet. 2016;99(4):877–885. doi:10.1016/j.ajhg.2016.08.016

33. Cheng J, Novati G, Pan J, et al. Accurate proteome-wide missense variant effect prediction with AlphaMissense. Science. 2023;381(6664):eadg7492. doi:10.1126/science.adg7492

34. Hopkins JJ, Wakeling MN, Johnson MB, Flanagan SE, Laver TW. REVEL Is Better at Predicting Pathogenicity of Loss-of-Function than Gain-of-Function Variants. Chen JM, ed. Hum Mutat. 2023;2023:1–6. doi:10.1155/2023/8857940

35. Cipriani V, Pontikos N, Arno G, et al. An Improved Phenotype-Driven Tool for Rare Mendelian Variant Prioritization: Benchmarking Exomiser on Real Patient Whole-Exome Data. Genes. 2020;11(4):460. doi:10.3390/genes11040460

36. Blasco M, Quiroga B, García-Aznar JM, et al. Genetic Characterization of Kidney Failure of Unknown Etiology in Spain: Findings From the GENSEN Study. Am J Kidney Dis Off J Natl Kidney Found. 2024;84(6):719–730.e1. doi:10.1053/j.ajkd.2024.04.021

37. Schwotzer N, Fakhouri F, Martins PV, et al. Hot Spot of Complement Factor I Rare Variant p.Ile357Met in Patients With Hemolytic Uremic Syndrome. Am J Kidney Dis. 2024;84(2):244–249. doi:10.1053/j.ajkd.2023.12.021

38. Timmermans SAMEG, van Doorn DPC, van Paassen P. Rare Variants in Complement Genes May Not Be That Rare After All. Kidney Int Rep. 2023;8(10):1911–1913. doi:10.1016/j.ekir.2023.08.020

39. Molinari E, Decker E, Mabillard H, et al. Human urine-derived renal epithelial cells provide insights into kidney-specific alternate splicing variants. Eur J Hum Genet EJHG. 2018;26(12):1791–1796. doi:10.1038/s41431-018-0212-5

40. O’Brien TD, Campbell NE, Potter AB, Letaw JH, Kulkarni A, Richards CS. Artificial intelligence (AI)-assisted exome reanalysis greatly aids in the identification of new positive cases and reduces analysis time in a clinical diagnostic laboratory. Genet Med. 2022;24(1):192–200.

