## Supplementary Table 1 for "Clinical Evaluation of an AI System for Streamlined Variant Interpretation in Genetic Testing" (from another version of this preprint: version 3, 2025-05-22)

### Supplemental Table 1

| **Characteristic** | **Overall**<br>N = 966 |
|---|---|
| Age, years | 45 (15) |
| Gender, male | 573 (69%) |
| Self-declared ancestry | |
| Europe | 344 (36%) |
| North Africa | 165 (17%) |
| Sub-Saharan Africa | 277 (29%) |
| Asian | 44 (5%) |
| Others or unknown | 136 (14%) |
| Diagnosed with exome, positive | 196 (20%) |
| Kidney biopsy performed | 346 (36%) |
| First-degree history of CKD | 311 (32%) |
| Mother | 99 (10%) |
| Father | 87 (9%) |
| Brother | 99 (10%) |
| Sister | 69 (7%) |
| Child | 27 (3%) |
| First or second-degree history of CKD | 409 (42%) |
| CKD stage at the time of ES | |
| 1 | 137 (16%) |
| 2 | 99 (11%) |
| 3 | 205 (24%) |
| 4 | 133 (15%) |
| 5 | 229 (26%) |
| Unknown | 63 (7%) |

**Supplemental Table 1. Characteristics of the patients included in the nephrology cohort used to evaluate DiagAI**. For continuous variables, we provide mean and standard deviation in parentheses. For discrete ones, we provide counts and proportions in parentheses. CKD: Chronic Kidney Disease, ES: Exome Sequencing.
